# Early life factors associated with childhood trajectories of violence among the Birth to Twenty-Plus Cohort in Soweto, South Africa

**DOI:** 10.1101/2023.10.27.23297689

**Authors:** Lilian Muchai, Sara Naicker, Juliana Kagura

## Abstract

Violence against children (VAC) has devastating and long-term negative consequences on individuals’ and society’s health, social and economic well-being. There is limited research on the life course experience of VAC, especially in Africa. This study aimed to identify sub-groups of physical & sexual violence victimization patterns separately in childhood, and evaluate early life factors predicting violence trajectories. This study used data from ages 5 to 18 years from the ongoing prospective Birth to Twenty Plus cohort (Bt20+). Children with data on physical and sexual violence in at least 2-time points between 5 and 18 years were included in the analyses. Group-based trajectory modelling was employed to identify groups of children with similar patterns of violence over time, while multivariable logistic regression was used to identify early life factors associated with violence trajectory group membership. Separately, two trajectory groups of physical violence (adolescent limited (65.1%) and chronic increasing (34.9%)) and sexual violence (adolescent limited (74.1%) and late increasing (25.9%)) victimization were identified. Early life factors associated with a higher risk of chronic increasing trajectory group membership, after adjusting for covariates, were being male (aOR 1.67, 95% CI 1.31; 2.10) and having a mother with at least secondary education compared to higher education (aOR 1.73, 95% CI 1.08; 2.76). In addition, residing in middle, compared to low, socioeconomic households (aOR 0.68, 95% CI 0.50; 0.92) was protective against membership in this group. Residing in high compared to low socioeconomic households, was the only early life factor with marginally significant (aOR 0.63, 95% CI 0.42; 0.95) association with membership in the late-increasing sexual violence victimization trajectory group. In conclusion, children follow different violence victimization trajectories over childhood. Identifying early life factors predicting violence trajectories provides key prevention intervention areas that can mitigate children’s violence experience.

## Introduction

Globally, more than half of children between the ages of 2 and 17 years reported having experienced some type of violence each year, in a systematic review on the global prevalence of childhood violence, which included reports published between the years 2000 and 2013 [1]. Africa bears the biggest brunt of violence against children (VAC), with the prevalence of physical violence at 60% and 51% for girls and boys respectively [2]. Violence is endemic to South Africa, particularly associated with structural and systemic violence originating from the Apartheid period. A nationally representative survey among children 15–17 years of age reported that almost 20% of children had experienced some form of sexual abuse, 21.3% experienced neglect, and over 30% and 16% experienced physical and emotional abuse respectively [3]. Children are likely to experience different forms of violence at different stages of their lives, as their environments and interactions change [4]. This ranges from violent discipline during early childhood by caregivers, bullying by peers at school, sexual violence victimization and intimate partner violence during the adolescent period – although the boundaries of these timeframes are porous [5,6]. A study conducted among children followed up from birth to 22 years in Soweto, found that 36% of children were exposed to violence at home, in the community, at school, peer violence and were victims of interpersonal violence [7].

Despite mounting evidence of determinants and the burden of VAC, few studies have demonstrated the effects of life course exposure to multiple forms of violence, and even fewer studies have identified the different clusters of violence patterns (trajectories) over time. Previous studies, carried out in developed countries have demonstrated heterogeneity in the number and shapes of trajectories of physical and/ or sexual violence victimization, with two [8,9] three [8,10], four [11] and five [12] trajectory groups identified. The majority of these studies assess trajectories of violence victimization as predictors of subsequent physical and mental health outcomes in adulthood, rather than the identification of risk factors for trajectory group membership. Most studies assessing risk factors for violence trajectory group membership in childhood tend to focus on violence aggression or perpetration. However, a significant overlap between victimization and perpetration experiences documented in research [13–16], show that victims and perpetrators share similar contextual features that place them at an increased risk for violence victimization and perpetration.

Several demographic and early life factors have been associated with violence trajectories. Being male, residing in households with low socioeconomic status, early motherhood, having more than one child in the household, single parenthood and low parental education were some of the risk factors identified for membership in high or life course persistent physical violence victimization or aggression trajectory groups [11,17–19]. From cross-sectional analyses conducted in both developing and developed countries, early life factors predicting violence victimization include, household crowding [20], paternal absence [21] and mother’s prior history of violence [22,23]. Both low [24] and high birth weight [25,26], and catch-up growth (weight and length) in infancy (0-2 years) are documented to predict future overweight and obesity [27,28], which are risk factors for violence victimization [29–31].

There is a dearth of studies conducted in Africa assessing factors associated with childhood trajectories of violence due to a lack of longitudinal data on violence. This limits the ability to assess the developmental course of violence and the heterogeneity of violence trajectories in populations outside of the Global North. This is paramount towards the identification, development and implementation of context-specific prevention strategies prior to the child’s exposure or experience of violence. The current study aims to generate physical and sexual violence victimization trajectories across childhood and assess how factors in early life predict membership into specific violence trajectories.

## Materials and methods

### Study setting and site

The primary study (Birth to Twenty plus study, Bt20+) is an ongoing prospective birth cohort study that follows singleton children born between April 23 to June 8, 1990 in public health facilities in Soweto, South Africa. Soweto is a densely populated suburb located within the Johannesburg metropolitan municipality in Gauteng province in South Africa. This primary study included pregnant women attending public antenatal clinics with expected delivery dates within the above-mentioned dates, and later continued residence of the mother and the baby within the study area during the child’s first six months of life. Data collected in the Bt20+ study is multidisciplinary, tracking infant, child, adolescent and now adult physical, social and psychosocial well-being [32]. Of 3,273 children enrolled at the beginning of the study, follow-up has been conducted bi-annually through home, study sites or through school surveys. Full descriptions of the Bt20+ cohort, its attrition and methods have been published elsewhere [32,33].

### Study design

This study involved secondary data analysis of the prospective Bt20+ cohort study. Only data from birth until 18 years covering (1990 – 2018) from the primary study was included in this secondary study.

### Study population and sample

The study population included all children from birth to 18 years from the primary ongoing prospective Bt20+ cohort study. This cohort of children is almost representative of the South African population, with a higher percentage of black Africans. However, there was a slight under-representation of white families [32].

From the 19 waves of data collection between birth to 18 years, there were only four time points, year 5, 11, 15 and 18, that had sufficient data on physical and sexual violence victimization. Data from data collection waves where physical and sexual violence data was combined were excluded from the analyses since trajectories were generated separately for physical & sexual violence victimization. Children in the study population with data on physical and sexual violence in at least 2 of the 4-time points were included in this study.

### Study variables

#### Outcome variable

Two outcome variables namely; physical and sexual violence victimization were considered for this study. For the early years, from age 5 to 10 years, the primary caregiver reported on the experiences of the participant, from age 11 onwards, experiences were self-reported.

Physical violence victimization was coded as a binary variable (yes/no) generated from questions on any experience of physical violence in the previous 12 months based on the following 4 categories: at home, at school, in the neighbourhood and at work (only year 18). The questions separately assessed for each of the four categories above included: have you been (a) physically hurt; (b) hit;(c) kicked; (d) badly beaten up; (e) attacked with a knife or sharp object. Responses to each of these questions were never, once or twice, few times and many times. The responses were recoded to a dichotomous variable with responses no, coded ‘0’ (never) and yes, coded ‘1’ (once or twice, few times and many times combined). A positive yes to any of the 5 questions for each of the 4 categories mentioned above was coded as having been a victim of physical violence (yes) for that specific time point of data collection.

Sexual violence victimization similar to the first outcome above was coded as a binary outcome of sexual violence experience in the preceding 12 months based on the categories (at home, at school, in the neighbourhood and at work) for questions a and b below. Questions c to g where not based on the specified categories. Sexual violence in the primary study was defined as unwanted sexual experiences. Questions included: have you been (a) sexually assaulted/ attacked (b) sexually harassed (c) wanted to engage in oral sex (d) wanted to have oral sex last month (e) wanted to have sex (f)wanted to have sex last month (g)wanted to engage in heavy petting or foreplay. Responses to questions a and b were never, once or twice, a few times or many times. These were recoded to no, coded ‘0’ (never) and yes, coded ‘1’ (once or twice, a few times and many times). Yes or no responses were recorded for questions c, d, e, f and g. Questions c to g were limited to children who had engaged in sex. These questions were recoded, where a no response, meaning for example that the individual did not want to engage in sex was recoded yes, coded ‘1’ to having been sexually assaulted. A yes response to question c, d, e, f and g was recoded no, coded ‘0’, to mean that the child had not been sexually assaulted. A positive yes to any of the questions above was coded as having been a victim of sexual violence (yes).

For details on the proportions of physical and sexual violence across the given time points please refer to supplementary table 1

#### Risk factors of interest

Early life factors measured between birth and 5 years associated with violence experience were selected based on aprior knowledge and literature, as well as data availability. These were selected as correlates for physical and sexual violence victimization trajectories and grouped into individual and family level factors.

##### Individual / Child-level factors

Birthweight was obtained from birth notifications and categorized as low birthweight (less than 2,500 grams) or normal birthweight (2,500 grams or higher). Relative weight gain and relative height gain were derived as standardized residuals computed from sex-specific linear regression of these growth changes measured between infancy (0-2) and early childhood (2-5 years).

##### Family level factors

Household socioeconomic status was based on total household asset scores, grouped into tertiles of low (coded ‘1’), middle (coded ‘2’) and high (coded ‘3’) socioeconomic status. Examples of some of the household assets included possession of a television, refrigerator, washing machine and a car. Household crowding was recorded as a ratio of persons per sleeping room, this was recoded to yes (‘1’) and no (‘0’). Household crowding was defined as households whose ratio was equal to or above the total sample household crowding ratio mean. Maternal age was recorded at birth and grouped for analysis - 24 years and below (coded ‘0’), between 25 and 34 years (coded ‘1’) and above 35 years (coded ‘2’) at the time of birth. Parity was coded as only child from mother (coded ‘0’) or mother had more than one child (coded ‘1’). Maternal and paternal education was categorized as primary school and below (coded ‘0’), secondary school (coded ‘1’), and post-school training (diploma, bachelors, masters or doctoral degrees, coded ‘2’). Marital status was recorded as married, living together, widowed/ divorced or single. This was recoded to married (married & living together, coded ‘1’) and single (widowed/divorced and single, coded ‘0’). Father’s presence in the household was coded as yes (‘1’) or no (‘0’). Maternal prior violence experience was derived using data from 2 variables. Any positive (yes) response on either mother’s prior violence in childhood or intimate partner violence during pregnancy was coded as mother having experienced violence (yes, coded ‘1’). Finally, mode of delivery obtained from birth notifications was recoded to normal vaginal delivery (‘0’) or assisted (caesarian section/ use of forceps or vacuum) delivery (‘1’).

### Data management and analysis

De-identified electronic data from the Birth to Twenty Plus study database was accessed on the 20^th^ of April 2023 following the ethical approval of this research. These data were imported into Stata 17 for cleaning and analysis. Only data on early life factors between the antenatal period to 5 years of age and data on physical and sexual violence victimization between ages 5 to 18 years were extracted from the primary Bt20+ database. The data was checked for missing and duplicate values, followed by recoding and generation of new variables. No duplicates were identified and the final study dataset was stored in a password-protected file for analyses.

#### Identification of trajectory groups

Group based trajectory modelling (GBTM) was used to identify clusters of children with similar violence victimization trajectories from age 5 to 18 years. GBTM is a semiparametric group strategy that applies finite mixture models to identify clusters of persons (trajectory groups) with similar developmental course of an outcome over time [34,35]. This technique assumes that the population is composed of distinct groups defined by their outcome progression over time and makes no assumptions about the population distribution of the trajectories [34]. Determination of the number of trajectory groups that best fit the data was based on the most parsimonious model determined by lowest Bayesian Information Criterion (BIC) and high entropy after comparison of two and three-group models. Linear, quadratic and cubic polynomial orders were tested to identify the polynomial order that best characterized the evolution of violence victimization over time. The logit function reflecting the Bernoulli distribution of the outcome was specified in all the models tested. The final model was selected based on theoretical plausibility, average posterior probabilities of trajectory group membership greater than the set threshold of 0.70, highest entropy and group membership greater than 10% [34]. All trajectory analyses were conducted using the stata plug-in Traj [36].

#### Early life factor correlates for trajectory membership

Descriptive analyses were conducted for the individual and family level early life factors disaggregated by sex. Once each of the children were assigned to trajectory groups to which each had maximum probability of group membership, a two steps process was used to identify early life factors associated with childhood trajectories of physical and sexual violence victimization. First, the independent association of each correlate to trajectory group membership was assessed using univariable logistic regression analysis. Odds ratios, 95% confidence intervals and p-values were reported. P-values <0.05 were considered statistically significant. Secondly, the selection of exposure variables for the multivariable model was based on documented evidence from existing literature of the associations between each of the correlates and violence victimization. Individual level factors were first added into the model, followed by family level factors. All correlates supported by literature and those with sufficient data were included in the final multivariable logistic regression model. Adjusted odds ratios (aOR), confidence intervals and p-values were reported following the multivariable analyses, with p-values <0.05 regarded as statistically significant. Post regression analyses conducted included a test for collinearity to assess if the explanatory variables in the model were inter-related. In addition, the Hosmer-Lemeshow goodness of fit test was conducted to test the final model fitness.

### Ethical considerations

Ethical approval for the primary Bt20+ study was obtained from the Human Ethics and Research Committee (HREC-Medical) of the University of Witwatersrand prior to the start of the study (certificate number M111182). Written informed consent was initially obtained from the primary caregivers of the participants and at the appropriate age both children and the caregivers then provided written informed consent. For this secondary data analyses, permission to use the data was granted by Development Pathways for Health Research Unit (DPHRU) responsible for the primary study and a memorandum of agreement signed. Ethical clearance for the current study was granted by HREC-Medical at the University of Witwatersrand (certificate number M230218). Confidentiality of data was maintained throughout the research period by use of anonymized data which was password protected and backed up for safety.

## Results

### Study characteristics

A total of 3,273 children were enrolled at birth, of these 2,057(62.9%) and 2,051(62.7%) children had data on physical & sexual violence victimization in at least 2 of the 4 time points of data collection, respectively. Experience of physical violence ranged from 14.0% at age 5, with a peak of 71.8% at age 15, that declined to 38.6% at 18 years of age. Sexual violence experiences ranged from 0.8% at 5 years to a peak of 28.9% at the age of 18 years (Supplementary table 1). For both physical and sexual violence victimization, there were no significant differences between included and excluded samples with reference to sex, maternal age at birth, father’s presence and birth weight. Compared to children excluded in this study, a greater proportion of children included in the study resided in households with middle-level socioeconomic status, lived in single parent households, with the Bt20+ child being an only child. In contrast, the included sample had a lower proportion of parents (both mothers and fathers) with post-school training compared to children in the excluded sample (Supplementary table 2 & 3). Covariates for physical and sexual violence victimization trajectories that were significantly different between included and excluded samples, were adjusted for in the multivariable analyses.

Similar proportions of children stratified by gender were reported among children included in the physical violence victimization and sexual violence victimization trajectory analyses (Table 1). Among the 2057 children included in the physical violence and 2051 children included in the sexual violence victimization trajectory analyses, there was a higher proportion of girls (52.5% physical violence; 52.4% sexual violence) than boys. More girls (12.7%) than boys (9.7%) were born with low birth weight. In both analyses, there was a decline in infant weight and height during infancy (0-2 years) for both girls and boys. During early childhood (2-5 years), a slight increase in weight was observed among boys, with no reported change in height for both boys and girls. Overall, a higher proportion of children were born in households with low socioeconomic status (65.0% physical violence; 64.8% sexual violence) and less than half of the children resided in crowded households (41.8% physical violence; 42.0% sexual violence). In both analyses, only 10.4% of the children were born to a mother above the age of 35 years, a higher proportion of these children had siblings (61.9% physical violence; 61.8% sexual violence) and approximately 62.0% were living in single parent households at birth. A higher proportion of fathers (18.8% physical violence; 18.7% sexual violence) than mothers (8.6%) had attained higher schooling after completion of secondary school and only 15.2% of the children indicated that the father was at home. For both analyses, less than a fifth of the mother’s indicated having previously experienced violence (18.4%) and most children were born through normal vaginal delivery (88.2%).

**Table 1.** Description of study sample characteristics stratified by sex.

|  | **Physical violence victimization** |  |  | **Sexual violence victimization** |  |  |
| --- | --- | --- | --- | --- | --- | --- |
| **Variables** | **Total N (%)** | **Female n (%)** | **Male n (%)** | **Total N (%)** | **Female n (%)** | **Male n (%)** |
| Total sample | 2057 | 1080 (52.5) | 977 (47.5) | 2051 | 1074 (52.4) | 977 (47.6) |
| **Outcome variables** |  |  |  |  |  |  |
| **Violence trajectories** |  |  |  |  |  |  |
| Adolescent limited | 1340 (65.1) | 770 (71.3) | 570 (53.8) | 1519 (74.6) | 801 (74.6) | 718 (73.5) |
| Increasing (chronic /late) | 717 (34.9) | 310 (28.7) | 407 (41.7) | 532 (25.9) | 273 (25.4) | 259 (26.5) |
| **Exposure variables** |  |  |  |  |  |  |
| **Individual level factors** |  |  |  |  |  |  |
| **Birth weight** |  |  |  |  |  |  |
| Low birth weight (<2500 grams) | 231 (11.3) | 137 (12.7) | 94 (9.7) | 230 (11.2) | 136 (12.7) | 94 (9.7) |
| Normal birth weight (≥ 2500 grams) | 1822 (88.8) | 942 (87.3) | 880 (90.4) | 1817 (88.8) | 937 (87.3) | 880 (90.4) |
| **Infant and child growth** |  |  |  |  |  |  |
| Relative weight gain 0-2 years <sup>a</sup> | 1401 | -0.01 (1.00) | -0.05 (1.00) | 1398 | -0.01 (1.00) | -0.05 (0.99) |
| Relative weight gain 2-5 years <sup>a</sup> | 1292 | -0.03 (1.03) | 0.02 (1.03) | 1291 | -0.03 (1.03) | 0.02 (1.03) |
| Relative height gain 0-2 years <sup>a</sup> | 1402 | -0.03 (0.99) | -0.06 (0.98) | 1399 | -0.03 (0.99) | -0.06 (0.98) |
| Relative height gain 2-5 years <sup>a</sup> | 1292 | 0.00 (0.97) | 0.00 (1.03) | 1291 | 0.00 (0.97) | 0.00 (1.03) |
| **Family level factors** |  |  |  |  |  |  |
| **Household socioeconomic status** |  |  |  |  |  |  |
| Low | 1217 (65.0) | 630 (64.3) | 587 (65.8) | 1212 (64.8) | 626 (64.1) | 586 (65.6) |
| Middle | 392 (20.9) | 200 (20.4) | 192 (21.5) | 392 (21.0) | 199 (20.4) | 193 (21.6) |
| High | 263 (14.1) | 150 (15.3) | 113 (12.7) | 266 (14.2) | 152 (15.6) | 114 (12.8) |
| **Household crowding** |  |  |  |  |  |  |
| Yes | 744 (41.8) | 378 (40.3) | 366 (43.5) | 745 (42.0) | 376 (40.3) | 369 (43.8) |
| No | 1034 (58.2) | 559 (59.7) | 475 (56.5) | 1030 (58.0) | 556 (59.7) | 474 (56.2) |
| **Maternal age at the time of birth** |  |  |  |  |  |  |
| ≤ 24 years | 962 (46.8) | 504 (46.8) | 458 (46.9) | 961 (46.9) | 502 (46.8) | 459 (47.0) |
| 25–34 years | 879 (42.8) | 466 (43.2) | 413 (42.3) | 875 (42.7) | 463 (43.2) | 412 (42.2) |
| **Variables** | **Total N (%)** | **Female n (%)** | **Male n (%)** | **Total N (%)** | **Female n (%)** | **Male n (%)** |
| ≥ 35 years | 214 (10.4) | 108 (10.0) | 106 (10.9) | 213 (10.4) | 107 (10.0) | 106 (10.9) |
| **Maternal parity** |  |  |  |  |  |  |
| 1 child | 783 (38.1) | 423 (39.2) | 360 (36.9) | 784 (38.2) | 423 (39.4) | 361 (37.0) |
| >1 child | 1274 (61.9) | 657 (60.8) | 617 (63.2) | 1267 (61.8) | 651 (60.6) | 616 (63.1) |
| **Marital status** |  |  |  |  |  |  |
| Married | 773 (37.8) | 394 (36.7) | 379 (39.1) | 773 (37.9) | 392 (36.7) | 381 (39.2) |
| Single | 1270 (62.2) | 679 (63.3) | 591 (60.9) | 1266 (62.1) | 676 (63.3) | 590 (60.8) |
| **Maternal education status** |  |  |  |  |  |  |
| Primary & below | 232 (12.3) | 124 (12.5) | 108 (12.1) | 233 (12.4) | 125 (12.6) | 108 (12.1) |
| Secondary | 1491 (79.1) | 786 (79.3) | 705 (78.9) | 1490 (79.1) | 785 (79.4) | 705 (78.8) |
| Post school training | 162 (8.6) | 81 (8.2) | 81 (9.1) | 161 (8.6) | 79 (8.0) | 82 (9.2) |
| **Paternal education status** |  |  |  |  |  |  |
| Primary & below | 121 (8.3) | 67 (8.8) | 54 (7.8) | 120 (8.3) | 66 (8.7) | 54 (7.8) |
| Secondary | 1061 (72.9) | 543 (71.4) | 518 (74.6) | 1060 (73.0) | 542 (71.6) | 518 (74.5) |
| Post-school training | 273 (18.8) | 151 (19.8) | 122 (17.6) | 272 (18.7) | 149 (19.7) | 123 (17.7) |
| **Father present** |  |  |  |  |  |  |
| Yes | 1568 (84.9) | 825 (85.0) | 743 (84.7) | 1564 (84.8) | 822 (84.9) | 742 (84.7) |
| No | 280 (15.2) | 146 (15.0) | 134 (15.3) | 280 (15.2) | 146 (15.1) | 134 (15.3) |
| **Maternal prior violence experience** |  |  |  |  |  |  |
| Yes | 184 (18.4) | 91 (17.3) | 93 (19.5) | 185 (18.4) | 92 (17.5) | 93 (19.5) |
| No | 818 (81.6) | 435 (82.7) | 383 (80.5) | 819 (81.6) | 434 (82.5) | 385 (80.5) |
| **Mode of delivery** |  |  |  |  |  |  |
| Vaginal delivery | 1012 (88.2) | 525 (88.8) | 487 (87.6) | 1006 (88.2) | 521 (88.8) | 485 (87.6) |
| Assisted delivery | 135 (11.8) | 66 (11.2) | 69 (12.4) | 135 (11.8) | 66 (11.2) | 69 (12.5) |

<sup>a</sup> Values are means (standard deviation)

### Trajectories of physical & sexual violence victimization

After the examination of different trajectory group numbers and polynomial functions, a two-group model provided the most parsimonious model that best described patterns for both physical & sexual violence victimization between 5 and 18 years of age. However, the shape or patterns of violence victimization differed between physical and sexual violence victimization trajectories.

The two group trajectories of physical violence victimization had an early onset of violence victimization. These groups were (a) adolescent limited (65.1%) characterized by gradual increase in violence victimization from age 5 to 15 years, after which there was a decrease in violence victimization beginning at age 15, (b) the chronic increasing group (34.9%) was characterized by a persistently increasing pattern of violence victimization at all time points (Fig 1). The two-group model selected had the lowest BIC value, with average posterior probabilities for group membership greater than 0.70 for the adolescent limited group (0.93) and the chronic increasing group (0.73). This model had medium entropy of 0.56 reported (Supplementary table 4).

**Fig 1.**
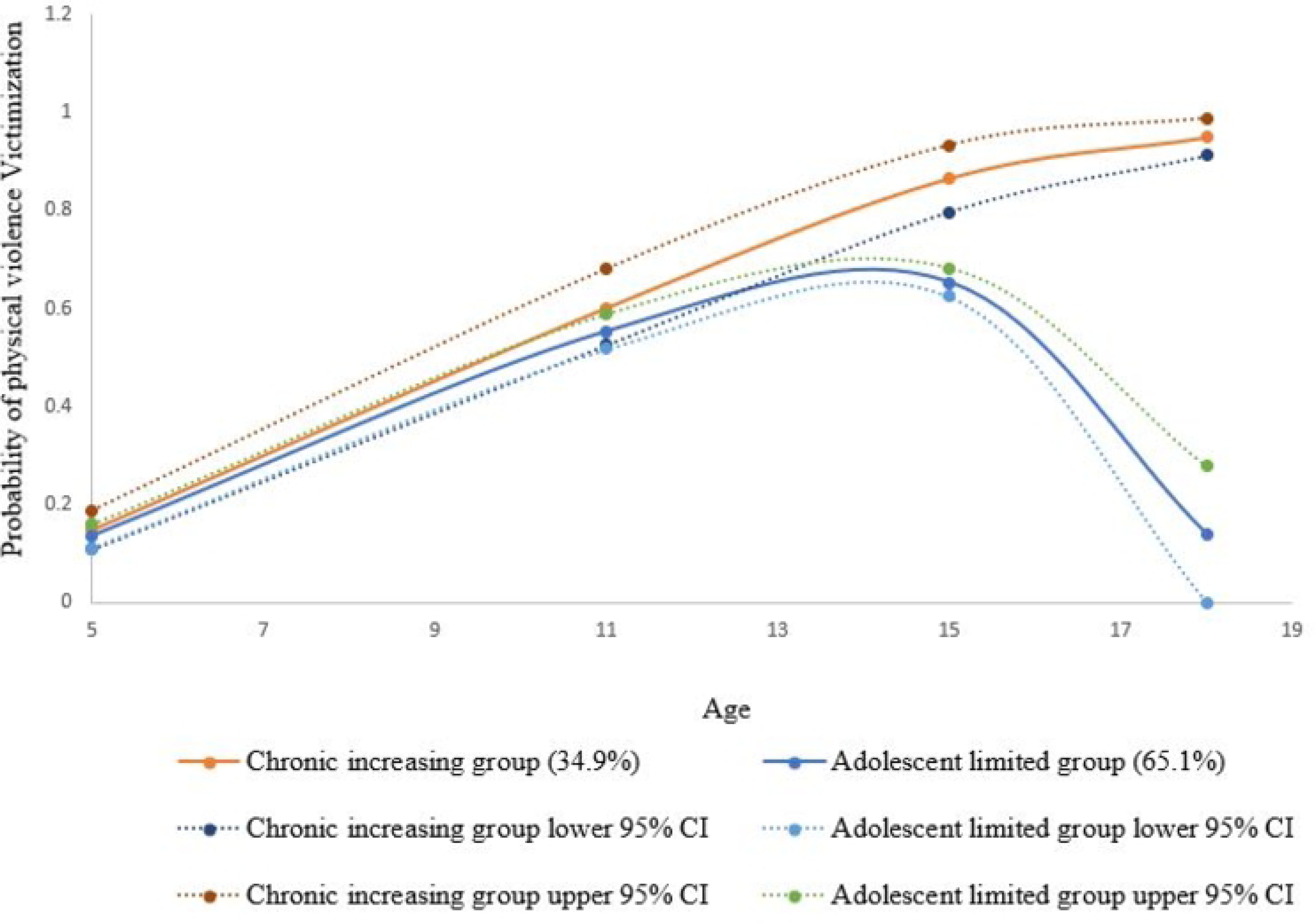
Physical violence victimization trajectories among children between 5 and 18 years of age. Figure 1 legend; Note; lines show estimated trajectories, points at each age represent the observed proportion of children between the age of 5 to 18 years reporting physical violence victimization according to the assigned physical violence trajectory group.

Figure 2 below shows the two-group trajectories of sexual violence victimization that were characterized by a late onset of sexual violence victimization. These groups were the (a) adolescent limited (74.1%) and (b) late increasing (25.9%) trajectory groups of sexual violence victimization. The adolescent limited group was characterized by low sexual violence victimization at the age of 5 years, whose onset was at age 11 with a peak at age 15. This was followed by a decrease in experience of sexual violence from age 15. Onset for the late increasing trajectory group was also at age 11, followed by a persistent increase in sexual violence victimization at the age of 15 continuing to 18 years. The model with a quadratic (2) and cubic (3) term provided the best description of sexual violence victimization trajectories. Although the 2-group model with quadratic terms had a slightly lower BIC compared to the model selected, the significantly lower entropy (0.44) for this model made it less desirable for describing sexual violence victimization patterns. The final model selected had average posterior probabilities of 0.85 and 1.00 for the adolescent limited and late increasing group, respectively, with an entropy of 0.57 (Supplementary table 4).

**Fig 2.**
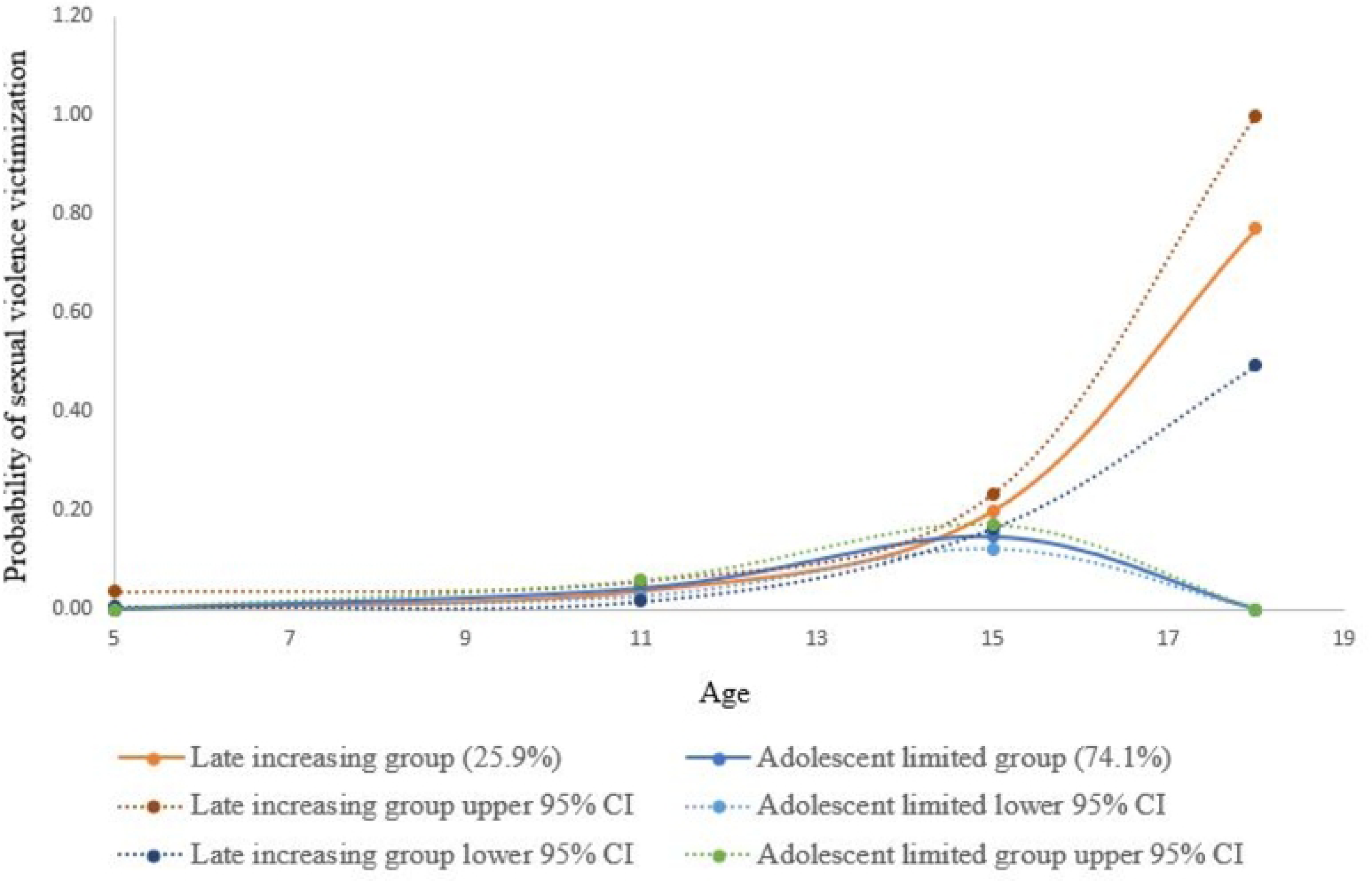
Sexual violence victimization trajectories among children between 5 and 18 years of age. Figure 2 legend; Note; lines show estimated trajectories, points at each age represent the observed proportion of children between the age of 5 to 18 years reporting sexual violence victimization according to the assigned sexual violence trajectory group.

### Early life factors associated with physical & sexual violence victimization trajectories

Table 2 and 3 show results of the independent and adjusted associations between the early life factors and physical and sexual violence victimization trajectory group membership, respectively. For physical violence victimization trajectories, individual and family level factors were significantly associated with trajectory group membership, both independently and after adjusting for other variables in the multivariable model. However, only family level factors significantly predicted sexual violence victimization trajectory group membership in the univariable, but not in the multivariable regression analyses.

**Table 2.** Univariable and multivariable logistic regression model showing early life factors associated with physical violence victimization trajectory group membership.

| Variables | OR (95% CI) | P-values | aOR (95% CI) | P-values |
| --- | --- | --- | --- | --- |
| **Individual level factors** |  |  |  |  |
| **Sex** |  | <0.001 |  | <0.001 |
| Female (ref) | 1.00 |  | 1.00 |  |
| Male | 1.77 (1.48; 2.13) |  | 1.67 (1.31; 2.10) |  |
| **Birth weight** |  | 0.416 |  | 0.479 |
| Normal (ref) | 1 |  | 1 |  |
| Low birth-weight | 1.12 (0.85; 1.49) |  | 1.15 (0.78; 1.68) |  |
| **Infants and child growth** |  |  |  |  |
| Relative weight gain 0-2 years | 1.14 (1.02; 1.27) | 0.023 |  |  |
| Relative weight gain 2-5 years | 0.98 (0.87; 1.09) | 0.664 |  |  |
| Relative height gain 0-2 years | 0.96 (0.86; 1.07) | 0.486 |  |  |
| Relative height gain 2-5 years | 0.88 (0.79; 0.99) | 0.035 |  |  |
| **Family level factors** |  |  |  |  |
| **Household socioeconomic status** |  | 0.006 |  | 0.034 |
| Low (ref) | 1.00 |  | 1.00 |  |
| Middle | 0.71 (0.55; 0.90) |  | 0.68 (0.50; 0.92) |  |
| High | 0.73 (0.55; 0.98) |  | 0.77 (0.54; 1.11) |  |
| **Household crowding** |  | 0.079 |  | 0.909 |
| Yes (ref) | 1.00 |  | 1.00 |  |
| No | 0.84 (0.69; 1.02) |  | 0.99 (0.77; 1.26) |  |
| **Maternal age** |  |  |  |  |
| ≤ 24 years (ref) | 1.00 | 0.975 | 1.00 | 0.465 |
| 25 – 34 years | 0.99 (0.82; 1.20) |  | 1.18 (0.87; 1.60) |  |
| ≥ 35 years | 1.03 (0.75; 1.40) |  | 1.29 (0.80; 2.08) |  |
| **Parity** |  | 0.395 |  | 0.601 |
| One child (ref) | 1.00 |  | 1.00 |  |
| more than one child | 1.08 (0.90; 1.31) |  | 0.92 (0.68; 1.25) |  |
| **Marital Status** |  | 0.096 |  | 0.238 |
| Married (ref) | 1.00 |  | 1.00 |  |
| Single | 1.17 (0.97; 1.42) |  | 1.18 (0.89; 1.57) |  |

| **Variables** | **OR (95% CI)** | **P-values** | **aOR (95% CI)** | **P-values** |
| --- | --- | --- | --- | --- |
| **Maternal education status** |  | 0.061 |  | 0.041 |
| Primary & below | 1.63 (1.05; 2.53) |  | 1.34 (0.74; 2.43) |  |
| Secondary | 1.52 (1.06; 2.19) |  | 1.73 (1.08; 2.76) |  |
| Post-school training (ref) | 1.00 |  | 1.00 |  |
| **Paternal education status** |  | 0.192 |  | 0.869 |
| Primary & below | 1.26 (0.80; 1.99) |  | 1.15 (0.66; 1.98) |  |
| Secondary | 1.31 (0.98; 1.74) |  | 1.08 (0.77; 1.51) |  |
| Post-school training (ref) | 1.00 |  | 1.00 |  |
| **Father present** |  | 0.473 |  | 0.624 |
| Yes (ref) | 1.00 |  | 1.00 |  |
| No | 1.10 (0.85; 1.43) |  | 1.10 (0.76; 1.59) |  |
| **Maternal prior violence experience** |  | 0.816 |  |  |
| Yes (ref) | 1.00 |  |  |  |
| No | 1.04 (0.74; 1.46) |  |  |  |
| **Mode of delivery** |  | 0.082 |  |  |
| Vaginal (ref) | 1.00 |  |  |  |
| Assisted | 0.70 (0.48; 1.04) |  |  |  |

OR: odds Ratio. aOR: adjusted Odds Ratio. Ref: reference group. Adolescent limited violence trajectory group acts as the outcome reference group. Goodness of fit of the model; $p=0.262$

Of the individual level factors, sex, relative infant weight gain and childhood height gain were independently associated with membership in the chronic increasing physical violence victimization trajectory group (table 2). From the univariable analysis, an increase in infant weight was associated with 14% greater odds of membership in the chronic increasing trajectory group (OR 1.14, 95% CI 1.02; 1.27). The results show that height gain during childhood was protective against membership in the chronic increasing trajectory group. An increase in relative height was associated with 22% lower odds of membership in the chronic increasing trajectory group (OR 0.88, 95% CI 0.79; 0.99). Relative growth variables during infancy and childhood could not be included in the multivariable model due to the high proportion of missing data. After adjusting for other variables in the multivariable model, sex was the only individual level factor that was significantly associated with membership in the chronic increasing trajectory group. Boys had 67% higher odds of membership in the chronic increasing trajectory group compared to girls (aOR 1.67, 95% CI 1.31; 2.10).

Household socioeconomic status was the only family level factor significantly associated with membership in the chronic increasing physical violence victimization trajectory group, in the unadjusted analysis. In addition, maternal education (p=0.061), household crowding (p=0.079) and mode of delivery (p=0.082) were marginally associated with chronic increasing trajectory group membership prior to adjusting for other factors. From the multivariable model, lower household socioeconomic status and lower maternal education were the only family level early life factors that predicted membership in the chronic increasing trajectory group, after adjusting for other variables in the model. Children from households within the middle socioeconomic status level had 32% lower odds of membership in the chronic increasing trajectory group compared to children residing in the low household socioeconomic status level (aOR 0.68, 95% CI 0.50; 0.92). Furthermore, children whose mothers had received at least some secondary education had 73% greater odds of chronic increasing trajectory group membership compared to mothers with post-school training (aOR 1.73, 95% CI 1.08; 2.76). Mother’s prior experience of violence and mode of delivery were not included in the multivariable analysis because of the high proportion of missing data for these two variables.

Results from table 3 demonstrate that only family level factors significantly predicted membership in the late increasing sexual violence victimization trajectory group, in the univariable regression analyses(p<0.05). Children from households with high socioeconomic status and no crowding had 42% (OR 0.58, 95% CI 0.41; 0.81) and 26% (OR 0.74, 95% CI 0.60; 0.92) lower odds of membership in the late increasing trajectory group compared to children from lower socioeconomic households and children from crowded households, respectively. Independently, the odds of membership in the late increasing trajectory group were greater with decreasing maternal (primary & below OR 2.56, 95% CI 1.55; 4.23, secondary OR 1.83, 95% CI 1.18; 2.83) and paternal (primary & below OR 1.70 95% CI 1.02; 2.83, secondary OR 1.60, 95% CI 1.19; 2.34) education compared to children from parents with post-school training. None of these factors remained statistically significant after adjusting for all other variables in the multivariable model. However, household socioeconomic status showed marginally significant (p=0.07) association with membership in the late increasing trajectory group in the multivariable analyses. Children residing in high socioeconomic households had 37% lower odds of membership in the late increasing trajectory group compared to children residing in households with low socioeconomic status (aOR 0.63, 95% CI 0.42; 0.95), after adjusting for covariates in the model. Due to the high proportion of missing data for growth variables, maternal prior violence experience and mode of delivery, these variables could not be included in the multivariable model.

**Table 3.** Univariable and multivariable logistic regression model showing early life factors associated with sexual violence victimization trajectory group membership.

| Variables | OR (95% CI) | P-values | aOR (95% CI) | P-values |
| --- | --- | --- | --- | --- |
| **Individual level factors** |  |  |  |  |
| **Sex** |  | 0.574 |  | 0.519 |
| Female (ref) | 1.00 |  | 1.00 |  |
| Male | 1.06 (0.87; 1.29) |  | 0.92 (0.71; 1.19) |  |
| **Birthweight** |  | 0.312 |  | 0.848 |
| Normal (ref) | 1.00 |  | 1.00 |  |
| Low birth-weight | 1.17 (0.86; 1.59) |  | 1.04 (0.69; 1.57) |  |
| **Infant and child growth** |  |  |  |  |
| Relative weight gain 0-2 years | 1.02 (0.90; 1.15) | 0.736 |  |  |
| Relative weight gain 2-5 years | 0.99 (0.88; 1.12) | 0.915 |  |  |
| Relative height gain 0-2 years | 0.95 (0.84; 1.07) | 0.393 |  |  |
| Relative height gain 2-5 years | 0.91 (0.80; 1.03) | 0.128 |  |  |
| **Family level factors** |  |  |  |  |
| **Household socioeconomic status** |  | <0.003 |  | 0.0671 |
| Low (ref) | 1.00 |  | 1.00 |  |
| Middle | 0.81 (0.62; 1.05) |  | 0.82 (0.59; 1.13) |  |
| High | 0.58 (0.41; 0.81) |  | 0.63 (0.42; 0.95) |  |
| **Household crowding** |  | 0.007 |  | 0.300 |
| Yes (ref) | 1.00 |  | 1.00 |  |
| No | 0.74 (0.60; 0.92) |  | 0.87 (0.66; 1.13) |  |
| **Maternal age** |  | 0.248 |  | 0.862 |
| ≤ 24 years (ref) | 1.00 |  | 1.00 |  |
| 25 – 34 years | 0.91 (0.74; 1.13) |  | 0.93 (0.67; 1.29) |  |
| ≥ 35 years | 1.20 (0.87; 1.67) |  | 1.02 (0.61; 1.72) |  |

| **Variables** | **OR (95% CI)** | **P-values** | **aOR (95% CI)** | **P-values** |
| --- | --- | --- | --- | --- |
| **Parity** |  | 0.332 |  | 0.721 |
| One child (ref) | 1.00 |  | 1.00 |  |
| more than one child | 1.11 (0.90; 1.36) |  | 1.06 (0.76; 1.47) |  |
| **Marital Status** |  | 0.761 |  | 0.768 |
| Married (ref) | 1.00 |  | 1.00 |  |
| Single | 1.03 (0.84; 1.27) |  | 1.05 (0.77; 1.42) |  |
| **Maternal education status** |  | 0.001 |  | 0.327 |
| Primary & below | 2.56 (1.55; 4.23) |  | 1.63 (0.85; 3.13) |  |
| Secondary | 1.83 (1.18; 2.83) |  | 1.31 (0.77; 2.23) |  |
| Post-school training (ref) | 1.00 |  | 1.00 |  |
| **Paternal education status** |  | 0.012 |  | 0.337 |
| Primary & below | 1.70 (1.02; 2.83) |  | 1.13 (0.62; 2.06) |  |
| Secondary | 1.67 (1.19; 2.34) |  | 1.32 (0.89; 1.94) |  |
| Post-school training (ref) | 1.00 |  | 1.00 |  |
| **Father present** |  | 0.741 |  | 0.225 |
| Yes (ref) | 1.00 |  | 1.00 |  |
| No | 0.95 (0.71; 1.27) |  | 0.77 (0.50; 1.18) |  |
| **Maternal prior violence experience** |  | 0.432 |  |  |
| Yes (ref) | 1.00 |  |  |  |
| No | 1.17 (0.80; 1.71) |  |  |  |
| **Mode of delivery** |  | 0.482 |  |  |
| Vaginal (ref) | 1.00 |  |  |  |
| Assisted | 0.86 (0.56; 1.31) |  |  |  |

OR: Odds Ratio. aOR: adjusted Odds Ratio. Ref: reference group. Adolescent limited violence trajectory group acts as the outcome reference group. Goodness of fit of the model; $p=0.520$

Results for the post regression goodness of fit analysis conducted after selection of final models for both physical (p=0.262) and sexual (p=0.520) violence victimization trajectories, did not provide evidence of a lack of model fitness. Low variance inflation factor (VIF) scores for both physical (mean VIF 1.2) and sexual (mean VIF 1.18) violence victimization from the multivariable analyses indicate that there was no multicollinearity among covariates included in each of the multivariable models.

## Discussion

The present study sought to examine the number and shapes of physical and sexual violence victimization trajectories between the ages of 5 and 18 years, as well as identify factors in early life associated with trajectory group membership. The results from the group-based trajectory modelling identified two trajectories of physical and two trajectories of sexual violence victimization, for the best description of physical violence victimization and sexual violence victimization patterns between the ages of 5 and 18 years. Overall, both the individual level (sex) and family level (household socioeconomic status and maternal education) factors were identified as important determinants of physical violence victimization trajectory group membership. However, vulnerability to increasing sexual violence victimization was largely determined by the family level factors (household socioeconomic status).

These findings show that less than a fifth of all the children in the sample experienced physical violence during early childhood. However, cases of physical violence victimization increased with age, as children engage with more environments outside the home. By mid-adolescence (15 years), almost three quarters of all the children were victims of physical violence. Physical violence victimization then begins to decline at 18 years. These findings are consistent with previous literature indicating that physical violence victimization is concentrated during childhood and early adolescent years and decreases with increasing age after mid-adolescence [37–39].

In accordance with literature, the majority of the children in the present study exhibited an increasing pattern of physical violence victimization as they grew older, with incidences of physical violence victimization primarily occurring between early childhood and mid-adolescence. However, for a third of the children, their experience of physical violence did not decrease in mid-adolescence, and gradually increased as they reached adulthood. Evidence of the presence of an adolescent limited and increasing physical violence trajectory group was reported by Semenza et al. [11], among individuals assessed across 4 data collection points between 12 and 34 years of age (wave 1 - 12-19 years, wave 2-13-20 years, wave 3 - 18-26 years and wave 4 - 24-34 years). Similar to our findings, the adolescent limited trajectory group was characterized by elevated levels of violence victimization in early adolescence that decreased rapidly as respondents transitioned to adulthood. In contrast, the increasing physical violence trajectory group was characterized by low experience of physical violence in adolescence which increased into adulthood. Two other trajectory groups, little to no victimization and high decreasing trajectory groups were identified by these authors [11].

Our results show that parent - reported cases of sexual violence victimization were less than 1% for children at 5 years. In general, incidence of sexual violence steadily increased with age during adolescence, and by the age of 18, nearly 1 in 3 children in the sample experienced sexual violence. The few to no parent-reported cases of sexual violence in early childhood may be a result of under-reporting or lack of knowledge of their child’s sexual experiences by parents or caregivers [40]. Other studies have documented a higher likelihood of sexual violence victimization among adolescent and adult samples compared to younger child samples [38,41]. A study among South African children between the ages of 10 and 17 years reported that, almost a quarter of child abuse perpetrators were intimate partners, a reason that can account for higher sexual abuse cases as children grow older and engage in intimate partner relationships [38].

For three quarters of the children, low sexual violence experiences were reported at 11 years, with incidences of sexual violence decreasing from 15 years of age. In contrast, a quarter of the children experience of sexual violence increased exponentially as they approached adulthood. Similar to our results, Jones et al. [9] generated two trajectory groups of sexual violence victimization assessed between the ages of 2 to 12 years. The two groups were; (a) no sexual violence victimization (b) a curvilinear pattern of sexual violence victimization characterized by low violence allegations at younger and older ages and a peak in sexual violence victimization between ages 4 to 8 years. The study recruited high risk children with data on alleged or substantiated allegations of child maltreatment or witnessing violence from child protective services records, a plausible reason for high sexual victimization experiences at younger ages [9].

Our findings show that individual level factors predicted membership in the chronic physical violence but not the late increasing sexual violence victimization trajectory group. For physical violence victimization, independent of other factors, boys were almost twice as likely as girls to be in the chronic increasing trajectory group. Similar findings have been demonstrated in other studies [2,11,37,42,43], and a possible reason for this is the fact that boys experience more conduct problems and exhibit greater externalizing behaviour compared to girls that places them at a heightened risk for both physical violence victimization and perpetration [18,44]. Although boys had a lower likelihood of membership in the late increasing sexual violence victimization group, this association was not significant. Contrary to this, many studies report significantly higher prevalence of sexual violence among girls than boys [38,41,45]. The lack of a significant difference in trajectory group membership between boys and girls may be explained by the use of self-completed questionnaires from the age of 11 years, that has been documented to contribute to higher disclosure on sexual violence experiences especially among boys [46].

From the univariable analyses increase in infant weight was a risk factor for chronic increasing physical violence trajectory group membership. In addition, being taller had a protective effect against membership in the persistently increasing physical violence victimization group. Rapid infant weight gain during infancy [24,28] and lower conditional height/ stunting during mid-childhood have been associated with overweight/ obesity in later childhood or adulthood [47,48], a risk factor for bullying and teasing in childhood [29,30]. None of the infant and child growth factors were associated with late increase sexual violence victimization group membership. Richter et al. [49] found conflicting results with stunted boys having a higher likelihood of sexual violence experience compared to boys with normal height for their age. Child growth factors could not be assessed further in adjusted analyses due to limited responses for these variables. Similar to findings from Startbuck et al. [50], birthweight was not significantly associated with physical and sexual violence victimization.

Family level factors independently, and when adjusted for other confounding variables, predicted membership in the increasing trajectory group for both physical (household socioeconomic status and maternal education) and sexual violence (household socioeconomic status only) victimization. Adjusting for other factors, higher household socioeconomic status was associated with lower risk of membership in the chronic increasing physical violence victimization trajectory group. This corresponds with evidence from other studies showing that physical violence experiences are higher among poorer households [43,51]. Higher socioeconomic status appeared to be protective against inclusion in the late increasing sexual violence victimization group. However, this association was not significant after other factors were taken into account. Richter et al. [49] found similar findings, with household socioeconomic status being only significantly associated with sexual abuse among boys prior to adjusting for other covariates.

Maternal education seems to be protective against chronic physical violence victimization trajectory group membership. Further evidence of this was reported by Semenza et al [11], who found that higher parental education was associated with a lower probability of membership in the increasing physical violence victimization compared to no victimization trajectory group. However, in our study higher maternal rather than paternal education was negatively associated with chronic physical violence victimization trajectory group membership. This could be explained by the study results by Sui et al. [13], which demonstrated that maternal presence in the household was associated with non-involvement in both violence victimization and perpetration.

From the independent analyses lower maternal and paternal education and household crowding were associated with a greater likelihood of membership in the late increasing sexual violence victimization trajectory group. This association, however, was not found to be significant after adjusting for other covariates in the model. Consistent with our findings, Ward et al. [46], found a marginal significant association in the univariable and not multivariable analyses between sharing a bedroom with more than one person and sexual abuse. Similar to our findings, marital status, maternal education and father’s presence were not found to be significantly associated with sexual abuse among boys in South Africa [49].

Interpretation of study findings should take into consideration the following limitations. First, presence of large gaps between data collection points when data on physical or sexual violence victimization could not be captured separately, may have affected the ability to discern more heterogeneity in violence victimization trajectories. Additional time points with violence victimization data in future research will provide a better characterization of violence patterns during childhood. High proportion of missing data limited the inclusion of growth variables into the multivariable model. Further research on the effect of infant catch-up and early childhood growth on violence victimization should be explored. The population under study represents an urban population, with a majority black population, and therefore results of this study should not be generalizable to the South African population. Finally, use of caregiver reports and self-reporting on sensitive issues such as sexual violence victimization may be subject to under-reporting or introduce social desirability bias. The large sample size and repeated collection of measures at different waves can reduce the magnitude of this bias.

Irrespective of the limitation mentioned above, several strengths of this study are worth mentioning. Studies identifying risk factors for violence victimization are based largely on cross-sectional or retrospective studies on lifetime childhood violence experiences. Results from these studies are subject to recall bias. This current study is based on a prospective study design with smaller recall period of 12 months limiting the magnitude of recall bias. In addition, the longitudinal nature of data collection allows for the characterization of violence victimization patterns. These violence patterns generated can further be assessed as predictors of later human capital outcomes in adulthood. Assessment of early life risk factors at the first years of life that predict violence trajectory group membership allow for the possibility of temporal association to be assessed.

In summary, this study demonstrates that children within the same environment or neighborhoods can follow different patterns of physical and sexual violence victimization from childhood to adolescence. The identification of early life factors associated with membership in these violence victimization groups, provides potential early target prevention areas that impede the risk of increasing violence victimization and also provides information on factors that can be encouraged to mitigate a child’s exposure and experience of violence. This study only identified individual and family early life factors associated with violence experience. Both these factors are heavily influenced by the community factors and structures set up in the society. There is a need for future research to include community and societal exposure factors when assessing risk factors for violence victimization trajectories.

## Data Availability

The Birth to Twenty-Plus Cohort data underlying the results presented in the study can be requested from the Birth to Thirty Executive Committee (https://bt30.org/data/). Alternatively, the authors can be contacted for a copy of the analytic dataset.

## Acknowledgements

The authors would like to thank the Birth to Twenty-Plus participants for their role in providing data for this work and also appreciate DSI-NRF Centre of Excellence in Human Development for granting access to use the Birth to Twenty-Plus Cohort data. We are grateful to the University of Witwatersrand HREC for provision of ethical clearance.

## Supporting information

**S1 Supplementary table 1. Proportions of physical and sexual violence victimization per age.**

**S2 Supplementary table 2. Characteristics of excluded and included sample for physical violence victimization trajectory analyses.**

**S3 Supplementary table 3. Characteristics of included and excluded sample for sexual violence victimization trajectory analyses.**

**Supplementary table 4. Physical and sexual violence victimization trajectory model selection and adequacy.**

**S5 Supplementary text. Sample Power calculation.**

## References

1. Hillis S, Mercy J, Amobi A, Kress H. Global Prevalence of Past-year Violence Against Children: A Systematic Review and Minimum Estimates. Pediatrics. 2016;137: e20154079. doi:10.1542/peds.2015-4079

2. Moody G, Cannings-John R, Hood K, Kemp A, Robling M. Establishing the international prevalence of self-reported child maltreatment: a systematic review by maltreatment type and gender. BMC Public Health. 2018;18: 1164. doi:10.1186/s12889-018-6044-y

3. Burton P, Ward CL, Artz L, Leoschut L. The Optimus Study on Child Abuse, Violence and Neglect in South Africa (2015) | PDF | Child Neglect | Child Abuse. Available from: https://www.scribd.com/document/314489410/First-ever-national-study-of-child-maltreatment

4. World Health Organization. INSPIRE Technical Package. [cited 6 Mar 2023]. Available from: https://www.who.int/teams/social-determinants-of-health/violence-prevention/inspire-technical-package

5. United Nations Children’s Fund (UNICEF). A Familiar Face: Violence in the lives of children and adolescents. In: UNICEF DATA. 1 Nov 2017 [cited 13 Oct 2022]. Available from: https://data.unicef.org/resources/a-familiar-face/

6. United Nations Educational, Scientific and Cultural Organization (UNESCO). Behind the numbers: ending school violence and bullying - UNESCO Digital Library. [cited 2 Mar 2023]. Available from: https://unesdoc.unesco.org/ark:/48223/pf0000366483

7. Richter LM, Mathews S, Kagura J, Nonterah E. A longitudinal perspective on violence in the lives of South African children from the Birth to Twenty Plus cohort study in Johannesburg-Soweto. S Afr Med J. 2018;108: 181. doi:10.7196/SAMJ.2018.v108i3.12661

8. Sumter SR, Baumgartner SE, Valkenburg PM, Peter J. Developmental Trajectories of Peer Victimization: Off-line and Online Experiences During Adolescence. J Adolesc Health. 2012;50: 607–613. doi: 10.1016/j.jadohealth.2011.10.251

9. Jones DJ, Runyan DK, Lewis T, Litrownik AJ, Black MM, Wiley T, et al. Trajectories of childhood sexual abuse and early adolescent HIV/AIDS risk behaviors: the role of other maltreatment, witnessed violence, and child gender. J Clin Child Adolesc Psychol Off J Soc Clin Child Adolesc Psychol Am Psychol Assoc Div 53. 2010;39: 667–680. doi:10.1080/15374416.2010.501286

10. Pahl K, Williams SZ, Lee JY, Joseph A, Blau C. Trajectories of Violent Victimization Predicting PTSD and Comorbidities among Urban Ethnic/Racial Minorities. J Consult Clin Psychol. 2020;88: 39–47. doi:10.1037/ccp0000449

11. Semenza DC, Testa A, Turanovic JJ. Trajectories of violent victimization over the life course: Implications for mental and physical health. Adv Life Course Res. 2021;50: 100436. doi: 10.1016/j.alcr.2021.100436

12. Tracy M, Salo M, Slopen N, Udo T, Appleton AA. Trajectories of childhood adversity and the risk of depression in young adulthood: Results from the Avon Longitudinal Study of Parents and Children. Depress Anxiety. 2019;36: 596–606. doi:10.1002/da.22887

13. Sui X, Massar K, Ruiter RAC, Reddy PS. Violence typologies and sociodemographic correlates in South African adolescents: a three-wave cross-sectional study. BMC Public Health. 2020;20: 221. doi:10.1186/s12889-020-8332-6

14. Maldonado-Molina MM, Jennings WG, Tobler AL, Piquero AR, Canino G. Assessing the victim-offender overlap among Puerto Rican youth. J Crim Justice. 2010;38: 1191–1201.

15. Jennings WG, Piquero AR, Reingle JM. On the overlap between victimization and offending: A review of the literature. Aggress Violent Behav. 2012;17: 16–26. doi: 10.1016/j.avb.2011.09.003

16. Miley LN, Fox B, Muniz CN, Perkins R, DeLisi M. Does childhood victimization predict specific adolescent offending? An analysis of generality versus specificity in the victim-offender overlap. Child Abuse Negl. 2020;101: 104328. doi: 10.1016/j.chiabu.2019.104328

17. Tremblay RE, Nagin DS, Séguin JR, Zoccolillo M, Zelazo PD, Boivin M, et al. Physical Aggression During Early Childhood: Trajectories and Predictors. Pediatrics. 2004;114: e43– e50. doi:10.1542/peds.114.1.e43

18. Gutman LM, Joshi H, Parsonage M, Schoon I. Gender-Specific Trajectories of Conduct Problems from Ages 3 to 11. J Abnorm Child Psychol. 2018;46: 1467–1480. doi:10.1007/s10802-017-0379-1

19. Nagin DS, Tremblay RE. Parental and Early Childhood Predictors of Persistent Physical Aggression in Boys from Kindergarten to High School. Arch Gen Psychiatry. 2001;58: 389–394. doi:10.1001/archpsyc.58.4.389

20. Gao Y, Mi X, Wang Y, Zou S, Zhou H. Association between Household Crowding and Violent Discipline and Neglect of Children: Analysis of Multiple Indicator Cluster Surveys in 26 Low- and Middle-Income Countries. Int J Environ Res Public Health. 2021;18: 1685. doi:10.3390/ijerph18041685

21. Palermo T, Pereira A, Neijhoft N, Bello G, Buluma R, Diem P, et al. Risk factors for childhood violence and polyvictimization: A cross-country analysis from three regions. Child Abuse Negl. 2019;88: 348–361. doi: 10.1016/j.chiabu.2018.10.012

22. Stith SM, Liu T, Davies LC, Boykin EL, Alder MC, Harris JM, et al. Risk factors in child maltreatment: A meta-analytic review of the literature. Aggress Violent Behav. 2009;14: 13–29. doi: 10.1016/j.avb.2006.03.006

23. Hayati Rezvan P, Tomlinson M, Christodoulou J, Almirol E, Stewart J, Gordon S, et al. Intimate Partner Violence and Food Insecurity Predict Early Behavior Problems Among South African Children over 5-years post-birth. Child Psychiatry Hum Dev. 2021;52: 409–419. doi:10.1007/s10578-020-01025-1

24. Salgin B, Norris SA, Prentice P, Pettifor JohnM, Richter LM, Ong KK, et al. Even transient rapid infancy weight gain is associated with higher BMI in young adults and earlier menarche. Int J Obes 2005. 2015;39: 939–944. doi:10.1038/ijo.2015.25

25. Evensen E, Emaus N, Kokkvoll A, Wilsgaard T, Furberg A-S, Skeie G. The relation between birthweight, childhood body mass index, and overweight and obesity in late adolescence: a longitudinal cohort study from Norway, The Tromsø Study, Fit Futures. BMJ Open. 2017;7: e015576. doi:10.1136/bmjopen-2016-015576

26. Kapral N, Miller SE, Scharf RJ, Gurka MJ, DeBoer MD. Associations between birthweight and overweight and obesity in school-age children. Pediatr Obes. 2018;13: 333–341. doi:10.1111/ijpo.12227

27. Ong KK, Ahmed ML, Emmett PM, Preece MA, Dunger DB. Association between postnatal catch-up growth and obesity in childhood: prospective cohort study. BMJ. 2000;320: 967–971. doi:10.1136/bmj.320.7240.967

28. Zheng M, Lamb KE, Grimes C, Laws R, Bolton K, Ong KK, et al. Rapid weight gain during infancy and subsequent adiposity: a systematic review and meta-analysis of evidence. Obes Rev Off J Int Assoc Study Obes. 2018;19: 321–332. doi:10.1111/obr.12632

29. Van Geel M, Vedder P, Tanilon J. Are overweight and obese youths more often bullied by their peers? A meta-analysis on the relation between weight status and bullying. Int J Obes. 2014;38: 1263–1267. doi:10.1038/ijo.2014.117

30. Haegele JA, Aigner C, Healy S. Impact of weight and disability status on bullying victimisation and perpetration among youth. J Paediatr Child Health. 2021;57: 383–387. doi:10.1111/jpc.15230

31. Janssen I, Craig WM, Boyce WF, Pickett W. Associations between overweight and obesity with bullying behaviors in school-aged children. Pediatrics. 2004;113: 1187–1194. doi:10.1542/peds.113.5.1187

32. Richter L, Norris S, Pettifor J, Yach D, Cameron N. Cohort Profile: Mandela’s children: The 1990 birth to twenty study in South Africa. Int J Epidemiol. 2007;36: 504–511. doi:10.1093/ije/dym016

33. Norris SA, Richter LM, Fleetwood SA. Panel studies in developing countries: case analysis of sample attrition over the past 16 years within the birth to twenty cohort in Johannesburg, South Africa. J Int Dev. 2007;19: 1143–1150. doi:10.1002/jid.1390

34. Nagin DS, Odgers CL. Group-Based Trajectory Modeling in Clinical Research. Annu Rev Clin Psychol. 2010;6: 109–138. doi: 10.1146/annurev.clinpsy.121208.131413

35. Nagin DS. Group-Based Trajectory Modeling: An Overview. Ann Nutr Metab. 2014;65: 205–210. doi:10.1159/000360229

36. traj: group-based modeling of longitudinal data. [cited 7 Aug 2023]. Available from: https://www.andrew.cmu.edu/user/bjones/

37. Macmillan R. Violence and the Life Course: The Consequences of Victimization for Personal and Social Development. Annu Rev Sociol. 2001;27: 1–22. doi: 10.1146/annurev.soc.27.1.1

38. Meinck F, Cluver LD, Boyes ME, Loening-Voysey H. Physical, emotional and sexual adolescent abuse victimisation in South Africa: prevalence, incidence, perpetrators and locations. J Epidemiol Community Health. 2016;70: 910–916. doi:10.1136/jech-2015-205860

39. Aboagye RG, Seidu A-A, Adu C, Cadri A, Mireku DO, Ahinkorah BO. Interpersonal violence among in-school adolescents in sub-Saharan Africa: Assessing the prevalence and predictors from the Global School-based health survey. SSM - Popul Health. 2021;16: 100929. doi:10.1016/j.ssmph.2021.100929

40. Pfeiffer L, Salvagni EP. Current view of sexual abuse in childhood and adolescence. J Pediatr (Rio J). 2005;81: 197–204. doi:10.2223/JPED.1408

41. Stoltenborgh M, van Ijzendoorn MH, Euser EM, Bakermans-Kranenburg MJ. A global perspective on child sexual abuse: meta-analysis of prevalence around the world. Child Maltreat. 2011;16: 79–101. doi:10.1177/1077559511403920

42. United Nations Children’s Fund (UNICEF). Hidden in plain sight: A statistical analysis of violence against children. [cited 13 Oct 2022]. Available from: https://www.unicef.org/documents/hidden-plain-sight-statistical-analysis-violence-against-children

43. Mahlangu P, Chirwa E, Machisa M, Sikweyiya Y, Shai N, Jewkes R. Prevalence and factors associated with experience of corporal punishment in public schools in South Africa. PLOS ONE. 2021;16: e0254503. doi: 10.1371/journal.pone.0254503

44. Richter LM, Ahun MN, Besharati S, Naicker SN, Orri M. Adolescent Mental Health Problems and Adult Human Capital: Findings from the South African Birth to Twenty Plus Cohort at 28 Years of Age. J Adolesc Health. 2021;69: 782–789. doi: 10.1016/j.jadohealth.2021.04.017

45. Cerna-Turoff I, Fang Z, Meierkord A, Wu Z, Yanguela J, Bangirana CA, et al. Factors Associated with violence against children in Low- and Middle-Income Countries: A Systematic Review and Meta-Regression of Nationally Representative Data. Trauma Violence Abuse. 2021;22: 219–232. doi:10.1177/1524838020985532

46. Ward CL, Artz L, Leoschut L, Kassanjee R, Burton P. Sexual violence against children in South Africa: a nationally representative cross-sectional study of prevalence and correlates. Lancet Glob Health. 2018;6: e460–e468. doi:10.1016/S2214-109X(18)30060-3

47. Bove I, Miranda T, Campoy C, Uauy R, Napol M. Stunting, overweight and child development impairment go hand in hand as key problems of early infancy: Uruguayan case. Early Hum Dev. 2012;88: 747–751. doi: 10.1016/j.earlhumdev.2012.04.002

48. Adair LS, Fall CHD, Osmond C, Stein AD, Martorell R, Ramirez-Zea M, et al. Associations of linear growth and relative weight gain during early life with adult health and human capital in countries of low and middle income: findings from five birth cohort studies. Lancet Lond Engl. 2013;382: 525–534. doi:10.1016/S0140-6736(13)60103-8

49. Richter LM, Mathews S, Nonterah E, Masilela L. A longitudinal perspective on boys as victims of childhood sexual abuse in South Africa: Consequences for adult mental health. Child Abuse Negl. 2018;84: 1–10. doi: 10.1016/j.chiabu.2018.07.016

50. Starbuck GW, Krantzler N, Forbes K, Barnes V. Child abuse and neglect on Oahu, Hawaii: description and analysis of four purported risk factors. J Dev Behav Pediatr. 1984;5: 55–59.

51. Meinck F, Cluver LD, Boyes ME, Ndhlovu LD. Risk and Protective Factors for Physical and Emotional Abuse Victimization amongst Vulnerable Children in South Africa: Physical and Emotional Child Abuse in South Africa. Child Abuse Rev. 2015;24: 182–197. doi:10.1002/car.2283

